# Associations of ambient air pollutants and meteorological factors with COVID-19 transmission in 31 Chinese provinces: A time-series study

**DOI:** 10.1101/2020.06.24.20138867

**Authors:** Han Cao, Bingxiao Li, Tianlun Gu, Xiaohui Liu, Kai Meng, Ling Zhang

## Abstract

**Background:** Evidence regarding the effects of ambient air pollutants and meteorological factors on COVID-19 transmission is limited.

**Objectives:** To explore the associations of air pollutants and meteorological factors with COVID-19 confirmed cases across 31 Chinese provinces during the outbreak period.

**Methods:** The number of COVID-19 confirmed cases, air pollutant concentrations and meteorological factors in 31 Chinese provinces from January 25 to February 29, 2020 were extracted from authoritative electronic databases. The associations were estimated for a single-day lag (lag0-lag6) as well as moving averages lag (lag01-lag05) using generalized additive mixed models (GAMMs), adjusted for time trends, day of the week, holidays and meteorological variables. Region-specific analyses and meta-analysis were conducted in five selected regions with diverse air pollution levels and weather conditions. Nonlinear exposure-response analyses were performed.

**Results:** We examined 77,578 COVID-19 confirmed cases across 31 Chinese provinces during the study period. An increase of each interquartile range in PM_2.5_, PM_10_, SO_2_, NO_2_, O_3_ and CO at lag4 corresponded to 1.40 (1.37-1.43), 1.35 (1.32-1.37), 1.01 (1.00-1.02), 1.08 (1.07-1.10), 1.28 (1.27-1.29) and 1.26 (1.24-1.28) odds ratios (ORs) of daily COVID-19 confirmed new cases, respectively. For 1 °C, 1% and 1 m/s increase in temperature, relative humidity and wind velocity, the ORs were 0.97 (0.97-0.98), 0.96 (0.96-0.97), and 0.94 (0.92-0.95), respectively. The estimates of PM_2.5_, PM_10_, NO_2_ and all meteorological factors remained statistically significant after meta-analysis for the five selected regions. The exposure-response relationships showed that higher concentrations of air pollutants and lower meteorological factors were associated with daily COVID-19 confirmed new cases increasing.

**Conclusions:** Higher air pollutant concentrations and lower temperature, relative humidity and wind velocity may favor COVID-19 transmission. As summer months are arriving in the Northern Hemisphere, the environmental factors and implementation of public health control measures may play an optimistic role in controlling COVID-19 epidemic.

## 1. Introduction

Since December 2019, coronavirus disease (COVID-19) caused by SARS-CoV2 has been epidemic in Wuhan, China (Li et al. 2020). An increasing number of COVID-19 confirmed cases have since been identified throughout China and the world, resulting in a declaration of a pandemic by the World Health Organization (WHO) due to human-to-human transmissibility and rapid spread across countries (Zhou et al. 2020). Environmental factors may influence the trends of infectious disease outbreaks by changing the host susceptibility and the survival time of viruses in vitro (Wu et al. 2016). Previous studies have found that air pollution, temperature and wind velocity were related to the outbreak of SARS (Cai et al. 2007; Cui et al. 2003; Kan et al. 2005; Tan et al. 2005), which has an 80% genetic sequence identity to COVID-19 (Lu et al. 2020). However, the associations of environmental factors with COVID-19 transmission remain incompletely understood.

Ambient air pollution exposures may play a role in respiratory diseases, such as asthma, influenza and SARS (Gehring et al. 2020; Kan et al. 2005; Liu et al. 2019). A time-series study has shown that particulate matter (PM) and nitrogen oxides (NO_x_) were positively associated with lower respiratory infection hospital admissions (Nhung et al. 2018). Kan et al. also found that 5-day moving average concentrations of PM with an aerodynamic diameter <10 μm (PM_10_) and nitrogen dioxide (NO_2_) corresponded to the increased daily mortality of SARS (Kan et al. 2005). However, the evidence of air pollutants and COVID-19 infection is limited with only two reported studies investigating the associations between them. Yao et al. found positive associations between NO_2_ levels and spread ability of COVID-19 across 63 Chinese cities (Yao et al. 2020a). Zhu et al. also reported that the number of COVID-19 confirmed cases was positively associated with PM_2.5_, PM_10_, NO_2_ and O_3_, but negatively associated with SO_2_ (Zhu et al. 2020). More epidemiological studies and laboratory evidence are needed to verify the current results.

Meteorological factors can impact the survival time of pathogens and change the host susceptibility, which further influence the trends of infectious disease outbreaks (Wu et al. 2016). Temperature and humidity affect influenza virus transmission and survival (Ianevski et al. 2019; Liu et al. 2018; Peci et al. 2019). Lowen et al. proposed that cold temperature and low relative humidity are favorable to the spread of influenza virus (Lowen et al. 2007). Limited studies have shown that temperature may change COVID-19 transmission significantly (Liu et al. 2020; Shi et al. 2020; Wang et al. 2020).

However, Yao et al. and Luo et al. found that there was no association of COVID-19 transmission with temperature and humidity, respectively (Luo et al. 2020; Yao et al. 2020b). Therefore, the effects of temperature and humidity on this novel pathogen are still unclear. In addition, wind velocity is another key factor affecting the pathogens of respiratory-borne diseases. A study on SARS data has suggested a negative correlation between daily average wind velocity and attack rate (Cai et al. 2007). However, evidence regarding the associations of wind velocity with COVID-19 infection remains sparse.

The present study aimed to assess the associations of ambient air pollutants and meteorological factors with COVID-19 confirmed cases across 31 Chinese provinces during the outbreak period (from January 25 to February 29, 2020) and to provide evidence for the further prevention and control of the COVID-19 epidemic.

## 2. Methods

### 2.1 Data collection

#### 2.1.1 Study population data

The study population was the daily confirmed new cases of COVID-19 in 31 Chinese provinces from January 25 to February 29, 2020. The number of cumulative confirmed cases was collected from reports released on the official websites of the National Health Commission (http://www.nhc.gov.cn/xcs/xxgzbd/gzbd_index.shtml), and then daily confirmed new cases were calculated and used in the present study.

Because the study population data was public data and was derived from official websites, there was no requirement for ethical review.

#### 2.1.2 Ambient air pollution data

The concentrations of PM_2.5_ (particulate matter <2.5 μm in aerodynamic diameter), PM_10_ (particulate matter <10 μm in aerodynamic diameter), nitrogen dioxide (NO_2_), sulfur dioxide (SO_2_), carbon monoxide (CO) and ozone (O_3_) were obtained from the China National Environmental Monitoring Centre (http://www.cnemc.cn/sssj/). Continuous hourly concentrations were gathered from January 25 to February 29, 2020. The daily average concentrations of PM_2.5_, PM_10_, SO_2_, NO_2_ and CO at each station were used only if >20 of the 24-hourly measurements were available. For O_3_, at least six hourly concentrations of O_3_ per day were needed to calculate the 8-hour average concentration of O_3_. The air pollutant concentrations were valid and were in accordance with the China Ambient Air Quality Standards (GB 3095-2012). Finally, daily pollutant concentrations of the province were averaged from all valid stations within it.

#### 2.1.3 Meteorological data

Daily mean ambient temperature, relative humidity and wind velocity from January 25 to February 29, 2020 (without missing data) were obtained from the National Meteorological Information Center (http://data.cma.cn/).

### 2.2 Statistical analysis

We characterized the distributions of all the covariates according to the mean and standard deviation (SD) or the median and interquartile range (IQR). The correlations between the concentrations of ambient air pollutants and meteorological factors during the study period were analyzed using Spearman rank correlation coefficients.

Generalized additive mixed models (GAMMs) were used to estimate the associations between air pollutants and daily COVID-19 confirmed new cases, in which days and provinces were treated as the first- and second-level units, respectively. Because the number of confirmed new cases approximately followed a Poisson distribution, a log-link function was used in GAMMs. Penalized cubic regression splines were used to capture time trends. We used the partial autocorrection function (PACF) of the residuals to determine the number of degrees of freedom for the spline function of time. Finally, we selected seven degrees of freedom in the spline function of time for all pollutants. In addition, we included the following potential confounders in the models: daily mean temperature, relative humidity, wind velocity, and categorical variables for day of the week and public holidays. We adjusted for meteorological factors averaged over the same day and the previous day as well as over the seven days preceding this period. The generalized cross-validation (GCV) values were used to select the best averaging period for meteorological factors. We used three degrees of freedom for the spline function of meteorological factors, which reportedly allowed adequate control for their effects on health outcomes (Samet et al. 2000). In all pollutant models, province was incorporated as a random effect, and the covariates were incorporated as fixed effects. The statistical analyses for the associations of meteorological factors were similar to those for air pollutants.

Regarding the levels of air pollutants and meteorological factors, the associations were estimated with different lag structures using a single-day lag from the current day up to the previous six days (lag0–lag6) as well as moving averages of the current and previous days (lag01–lag06). To facilitate comparisons, the associations of air pollutants and meteorological factors with daily confirmed new cases were reported as odds ratio (OR) with 95% confidence interval (95% CI) for an IQR increase in daily average concentrations of air pollutants as well as 1 °C, 1% and 1 m/s increase in daily mean temperature, relative humidity and wind velocity, respectively.

Considering the heterogeneity of 31 Chinese provinces, we conducted subgroup analyses to explore region-specific associations. From the north to south of China, five regions (i.e., Heilongjiang, Beijing, Hubei, Guangdong and Hainan) with sufficient sample size of COVID-19 confirmed cases were investigated due to their diversity of air pollution levels and climate conditions. The total effects of air pollutants and meteorological factors in five regions were then calculated through meta-analysis by fixed models or random effect models.

The nonlinear exposure-response relationships were examined because linearity assumption of air pollutants and meteorological factors with COVID-19 confirmed cases may not hold. Instead of linear parameters, we included a penalized cubic regression spline of air pollutants or meteorological factors with three degrees of freedom into the GAMMs. To determine the susceptible weather characteristics, we further investigated the three-dimensional exposure-response relationships of temperature, relative humidity and wind velocity by including an interaction term with two of them into the models.

To assess the robustness of the associations of air pollutants and meteorological factors with COVID-19 confirmed cases, we conducted sensitivity analyses using four degrees of freedom in meteorological factors and varying the level of smoothness of the time trend function (with six to eight degrees of freedom).

All of the statistical analyses were performed using R version 3.5.3 (https://www.r-project.org/). A *P*-value <0.05 was considered as statistically significant for a two-tailed test.

## 3. Results

### 3.1 Descriptive statistics

Descriptive statistics for daily number of COVID-19 confirmed new cases, air pollutant concentrations, and meteorological factors are summarized in **Table 1**. We examined 77,578 COVID-19 confirmed new cases (approximately 2155 cases per day) in 31 Chinese provinces from January 25 to February 29, 2020. The concentrations of PM_2.5_, PM_10_, SO_2_, NO_2_, O_3_ and CO varied greatly during the study period with medians (IQR) of 46.7 (14.6) μg/m^3^, 62.6 (17.6) μg/m^3^, 10.6 (2.0) μg/m^3^, 18.5 (4.1) μg/m^3^, 80.9 (14.3) μg/m^3^, and 0.8 (0.2) mg/m^3^, respectively. Meteorological factors ranged from 1.4 °C to 11.6 °C in temperature, from 48.7% to 78.8% in relative humidity, and from 2.1 m/s to 4.7 m/s in wind velocity. As shown in **Table 2**, the daily concentrations of air pollutants were strongly and positively correlated with each other with the exception of O_3_ and CO. Meteorological factors were moderately correlated with each other. In addition, wind velocity was negatively correlated with all air pollutants. The distribution of air pollutant concentrations and meteorological factors was diverse among the five selected regions (i.e., Heilongjiang, Beijing, Hubei, Guangdong and Hainan). For instance, daily average concentration of PM_2.5_ ranged from 13.6 μg/m^3^ in Hainan to 72.0 μg/m^3^ in Beijing, and daily mean temperature ranged from −12.1 °C in Heilongjiang to 19.9 °C in Hainan **(Supplementary Table A1)**.

**Table 1.** Descriptive statistics for daily COVID-19 confirmed new cases, air pollutant concentrations, and meteorological factors in 31 Chinese provinces from January 25 to February 29, 2020.

| Variable | Total | Mean | SD | Min | P25 | Median | P75 | Max | IQR |
| --- | --- | --- | --- | --- | --- | --- | --- | --- | --- |
| **Daily number of outcomes** |  |  |  |  |  |  |  |  |  |
| Confirmed cases, N | 77,578 | 2155 | 2467 | 126 | 658 | 1836 | 2692 | 14,893 | 2035 |
| **Daily air pollutant concentrations** |  |  |  |  |  |  |  |  |  |
| PM<sub>2.5</sub>, µg/m<sup>3</sup> | - | 46.1 | 13.7 | 16.1 | 38.0 | 46.7 | 52.7 | 85.6 | 14.6 |
| PM<sub>10</sub>, µg/m<sup>3</sup> | - | 61.9 | 14.4 | 32.1 | 53.9 | 62.6 | 71.5 | 101.7 | 17.6 |
| SO<sub>2</sub>, µg/m<sup>3</sup> | - | 10.7 | 1.9 | 7.1 | 9.7 | 10.6 | 11.7 | 17.3 | 2.0 |
| NO<sub>2</sub>, µg/m<sup>3</sup> | - | 18.0 | 3.5 | 9.8 | 16.0 | 18.5 | 20.0 | 24.9 | 4.1 |
| O<sub>3</sub>, µg/m<sup>3</sup> | - | 80.4 | 8.4 | 65.9 | 73.0 | 80.9 | 87.3 | 98.6 | 14.3 |
| CO, mg/m<sup>3</sup> | - | 0.8 | 0.1 | 0.5 | 0.8 | 0.8 | 0.9 | 1.0 | 0.2 |
| **Daily meteorological factors** |  |  |  |  |  |  |  |  |  |
| Temperature, °C | - | 5.7 | 3.1 | 1.4 | 2.9 | 4.7 | 8.4 | 11.6 | 5.5 |
| Relative humidity, % | - | 68.3 | 7.8 | 48.7 | 63.8 | 69.2 | 74.6 | 78.8 | 10.8 |
| Wind velocity, m/s | - | 2.8 | 0.6 | 2.1 | 2.4 | 2.7 | 3.0 | 4.7 | 0.6 |

Abbreviations: PM<sub>2.5</sub>, particulate matter <2.5 µm in aerodynamic diameter; PM<sub>10</sub>, particulate matter <10 µm in aerodynamic diameter; SO<sub>2</sub>, sulfur dioxide; NO<sub>2</sub>, nitrogen dioxide; CO, carbon monoxide; O<sub>3</sub>, ozone; SD, standardized deviation; Min, minimum; Max, maximum; P25, 25th percentile; P75, 75th percentile; IQR, interquartile range.

**Table 2.** Spearman rank correlation coefficients between daily air pollutant concentrations and meteorological factors.

| Variable | PM<sub>2.5</sub> | PM<sub>10</sub> | SO<sub>2</sub> | NO<sub>2</sub> | O<sub>3</sub> | CO | Temperature | Relative humidity | Wind velocity |
| --- | --- | --- | --- | --- | --- | --- | --- | --- | --- |
| PM<sub>2.5</sub> | 1.00 | 0.80<sup>a</sup> | 0.78<sup>a</sup> | 0.31 | 0.24 | 0.89<sup>a</sup> | -0.13 | 0.05 | -0.41<sup>b</sup> |
| PM<sub>10</sub> | - | 1.00 | 0.63<sup>a</sup> | 0.62<sup>a</sup> | 0.50<sup>b</sup> | 0.56<sup>a</sup> | 0.23 | -0.15 | -0.21 |
| SO<sub>2</sub> | - | - | 1.00 | 0.32 | 0.32 | 0.76<sup>a</sup> | -0.20 | -0.16 | -0.52<sup>b</sup> |
| NO<sub>2</sub> | - | - | - | 1.00 | 0.58<sup>a</sup> | 0.18 | 0.68<sup>a</sup> | -0.08 | -0.33<sup>b</sup> |
| O<sub>3</sub> | - | - | - | - | 1.00 | -0.04 | 0.30 | -0.78<sup>a</sup> | -0.24 |
| CO | - | - | - | - | - | 1.00 | -0.15 | 0.33<sup>b</sup> | -0.49<sup>b</sup> |
| Temperature | - | - | - | - | - | - | 1.00 | 0.12 | -0.08 |
| Relative humidity | - | - | - | - | - | - | - | 1.00 | 0.02 |
| Wind velocity | - | - | - | - | - | - | - | - | 1.00 |

<sup>a</sup> *P*-value <0.001.
<sup>b</sup> *P*-value <0.05.

The time series distribution of daily COVID-19 confirmed new cases, air pollutant concentrations and meteorological factors in 31 Chinese provinces are presented in **Figure 1**. The epidemic curve of COVID-19 daily confirmed new cases reached a peak twice (5 February and 13 February) during the study period with 3841 cases and 14,893 cases, respectively. The periodic peaks of PM_2.5_, PM_10_, SO_2_, NO_2_ and O_3_ concentrations were four days earlier than confirmed cases. The time series plots of temperature and relative humidity presented inverse trends compared to confirmed cases, especially after February 16, 2020.

**Figure 1.**
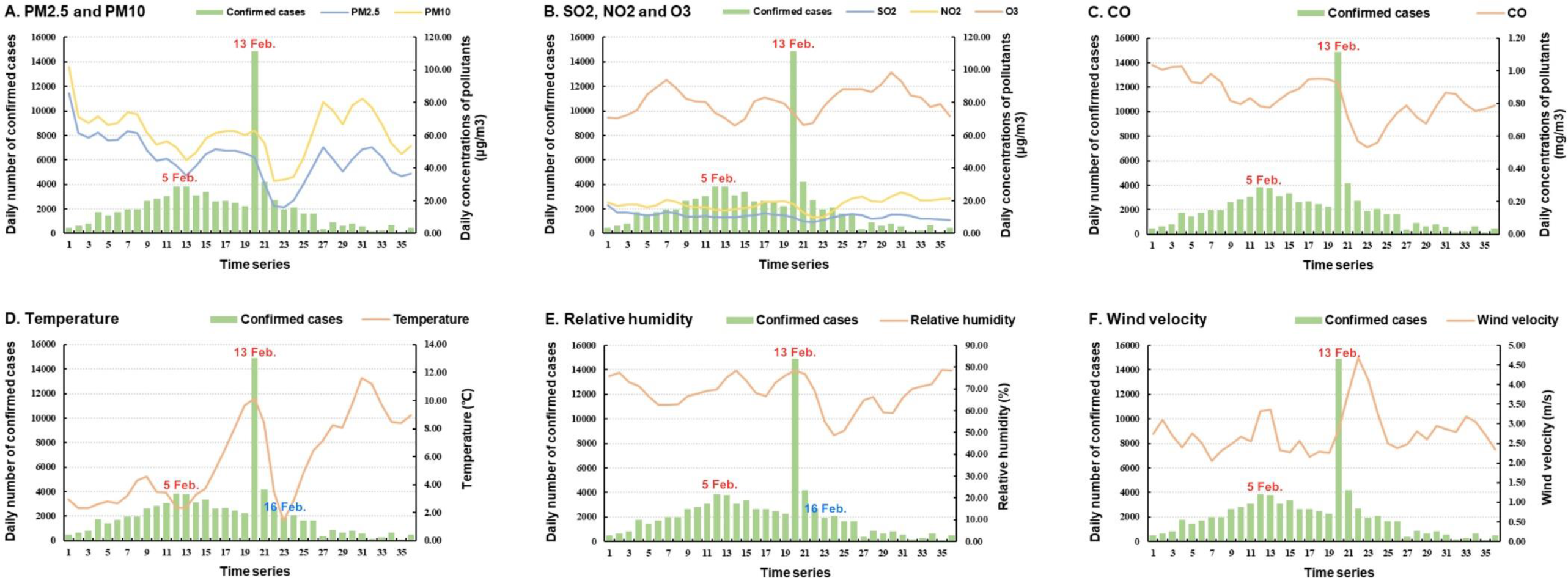
The time series distribution of daily COVID-19 confirmed new cases, air pollutant concentrations, and meteorological factors in 31 Chinese provinces from January 25 to February 29, 2020. Note: Columns: daily COVID-19 confirmed new cases. Curves: daily mean air pollutant concentrations and meteorological factors.

### 3.2 Associations of air pollutants and meteorological factors with COVID-19

The ORs of COVID-19 confirmed new cases per an IQR increase in the concentrations of air pollutants with different lag structures are presented in **Figure 2**. For the single-day lag, similar trends were observed in air pollutant estimates, reaching peaks at lag0 and lag4, except for SO_2_ (reaching peaks at lag1 and lag3) and O_3_ (reaching peaks at lag1 and lag4). The number of COVID-19 confirmed new cases was significantly positively associated with all pollutants at lag4, corresponding to 1.40 (1.37-1.43), 1.35 (1.32-1.37), 1.01 (1.00-1.02), 1.08 (1.07-1.10), 1.28 (1.27-1.29) and 1.26 (1.24-1.28) ORs per an IQR increase in the concentrations of PM_2.5_, PM_10_, SO_2_, NO_2_, O_3_ and CO, respectively. For the moving averages, positive significant associations were observed for the following parameters: PM_2.5_ at lag01 and lag04; PM_10_ at lag01, lag05 and lag06; SO_2_ at lag01-lag05; NO_2_ at lag01-lag03, lag05 and lag06; and O_3_ and CO at lag01-lag06.

**Figure 2.**
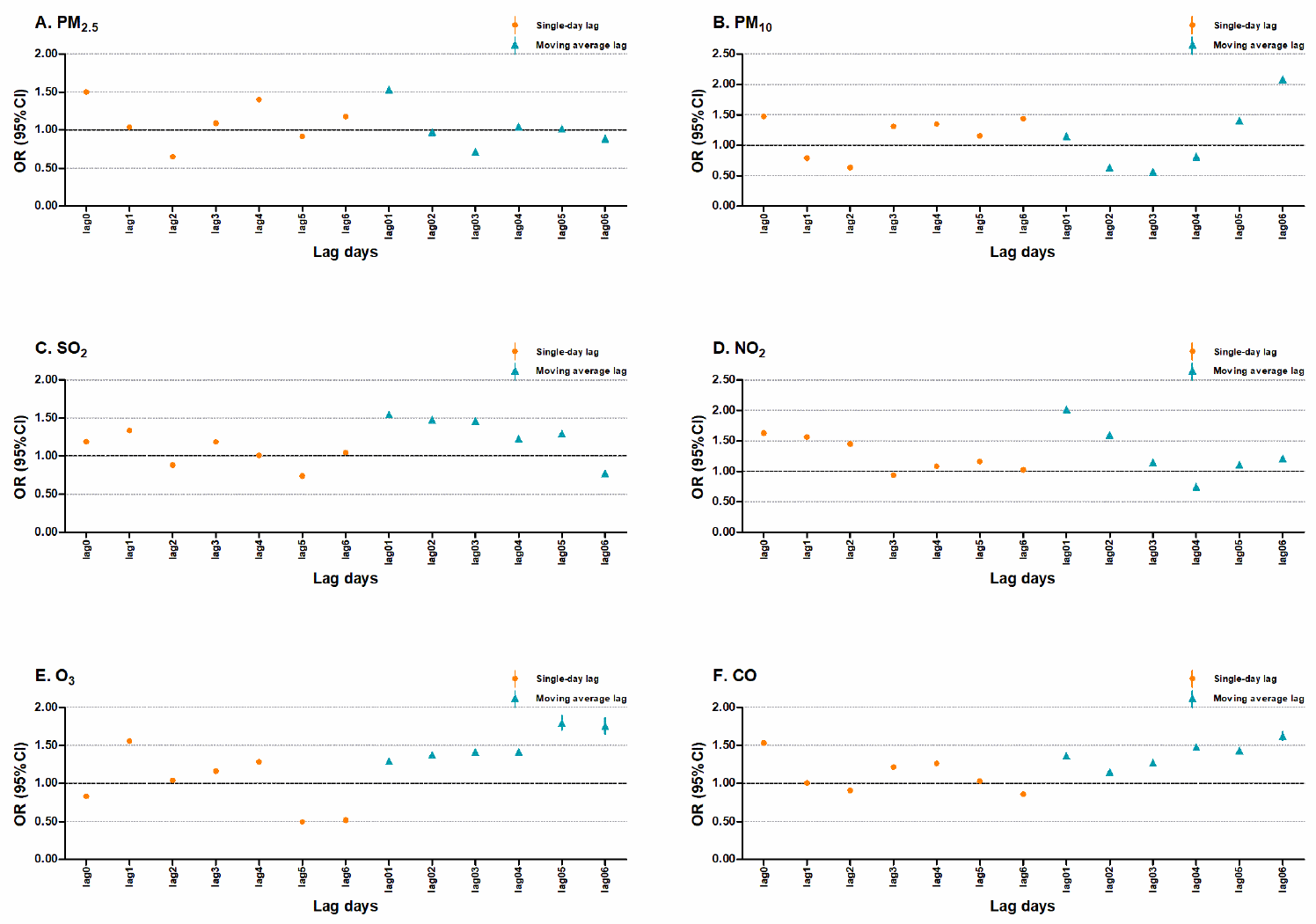
Odds ratios with 95% confidence intervals per an interquartile range increase in the concentrations of air pollutants using different lag structures for COVID-19 confirmed new cases in 31 Chinese provinces. Note: Orange circles: a single-day lag from the current day up to the previous six days (lag0-lag6). Blue triangles: moving averages of the current and previous days (lag01-lag06).

The associations between meteorological factors and COVID-19 confirmed new cases using different lag structures are shown in **Figure 3**. The negative significant associations were observed for all meteorological factors at both single-day and moving average lag structures with the following exceptions: temperature at lag0, lag1, lag01 and lag02; relative humidity at lag0, lag5, lag6, lag01, lag05 and lag06; and wind velocity at lag2 and lag3. Regarding the incubation period of COVID-19, we focused on the associations at the previous four days lag structure (lag4). The estimates per 1 °C, 1% and 1 m/s increase in temperature, relative humidity and wind velocity at lag4 were 0.97 (0.97-0.98), 0.96 (0.96-0.97), and 0.94 (0.92-0.95), respectively.

**Figure 3.**
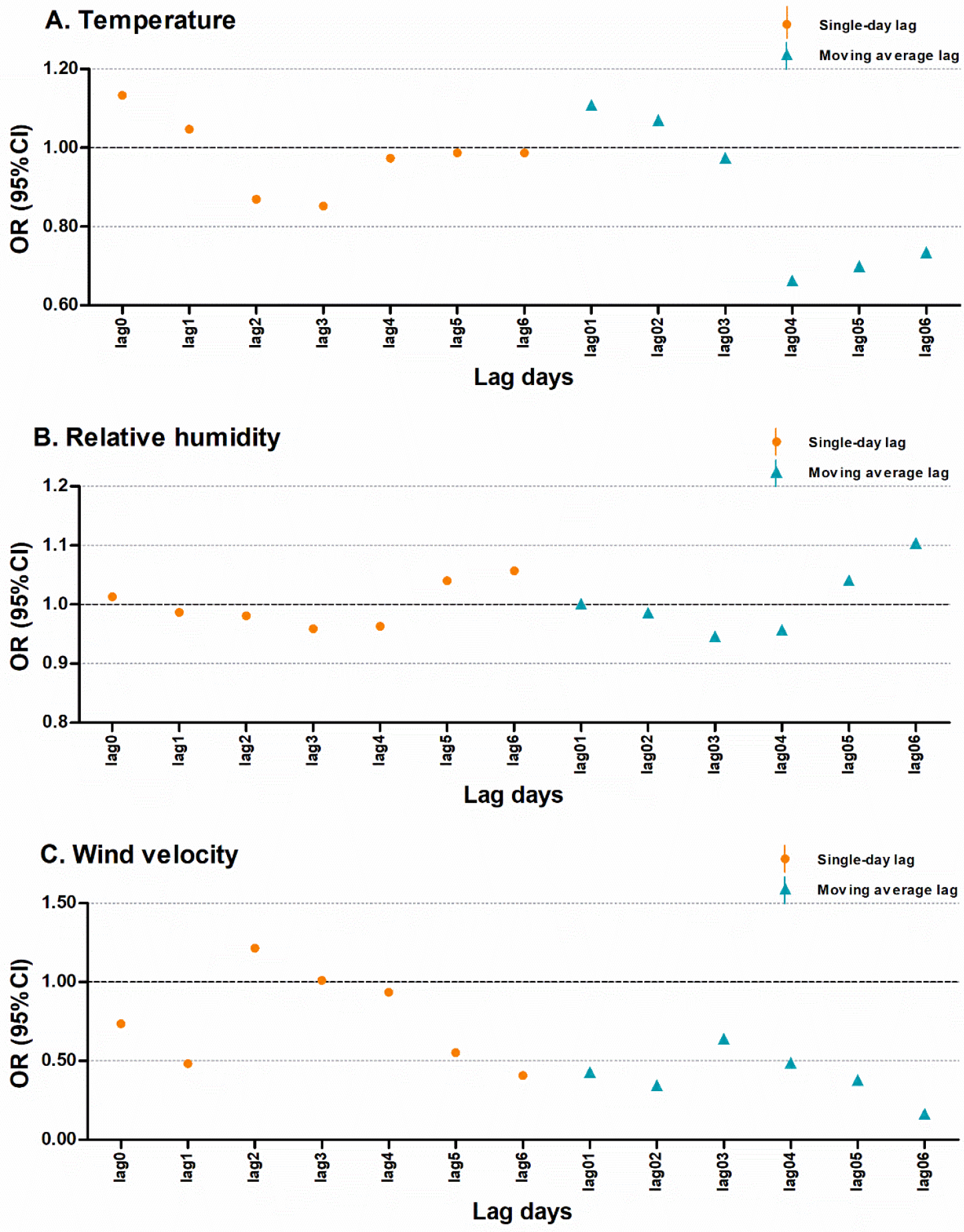
Odds ratios with 95% confidence intervals per 1 °C, 1% and 1 m/s increase in temperature, relative humidity and wind velocity using different lag structures for COVID-19 confirmed new cases in 31 Chinese provinces. Note: Orange circles: a single-day lag from the current day up to the previous six days (lag0-lag6). Blue triangles: moving averages of the current and previous days (lag01-lag06).

### 3.3 Region-specific associations and meta-analysis

Meta-analysis of the associations between air pollutant concentrations and COVID-19 confirmed new cases at lag4 among the five selected regions are presented in **Figure 4**. The number of confirmed new cases was significant positively associated with PM_2.5_, PM_10_, NO_2_, O_3_ and CO in both Heilongjiang and Hubei. Positive significant association of SO_2_ was only observed for Hubei. However, negative significant associations were observed for SO_2_ in both Beijing and Hainan as well as O_3_ in Beijing. The estimates of PM_2.5_ (1.13, 95% CI: 1.10-1.15), PM_10_ (1.20, 95% CI: 1.17-1.23) and NO_2_ (1.12, 95% CI: 1.11-1.13) remained significantly positive after meta-analysis for the five regions. In contrast, the estimates of SO_2_, O_3_ and CO lost statistical significance after meta-analysis.

**Figure 4.**
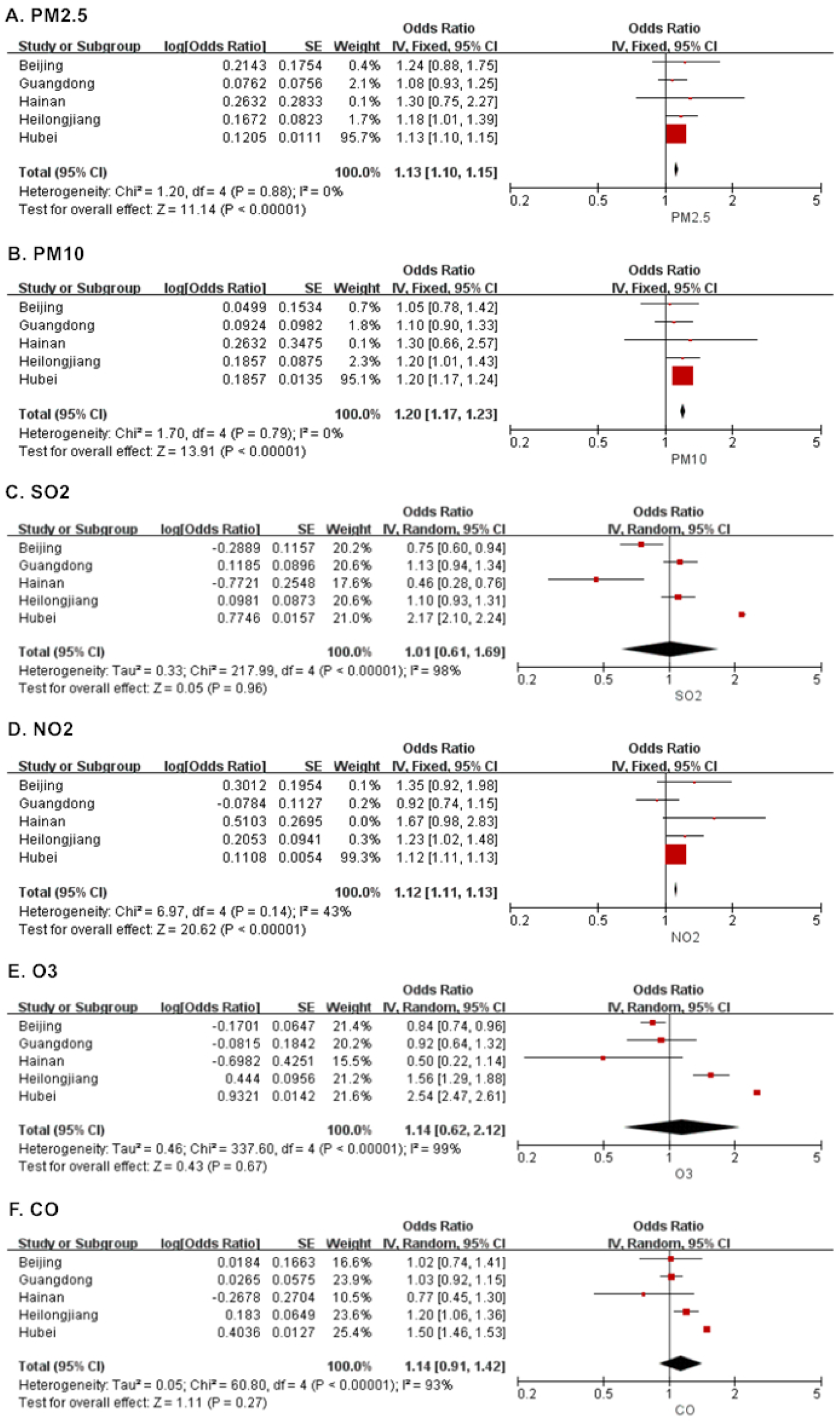
Meta-analysis of the associations between air pollutant concentrations and COVID-19 confirmed new cases at the previous four days lag structure (lag4) in the five selected regions.

Meta-analysis results of the ORs of COVID-19 confirmed new cases with meteorological factors at lag4 among the five regions are shown in **Figure 5**. The number of confirmed new cases was significantly negatively associated with temperature and relative humidity among Guangdong, Heilongjiang and Hubei. Negative associations were observed for wind velocity in Beijing, Guangdong, Heilongjiang and Hubei. However, the estimate of wind velocity with statistical significance were only examined in Hubei. After meta-analysis of the five regions, the estimates of temperature (0.95, 95% CI: 0.90-1.00), relative humidity (0.99, 95% CI: 0.98-1.00) and wind velocity (0.90, 95% CI: 0.82-0.98) remained significantly negative.

**Figure 5.**
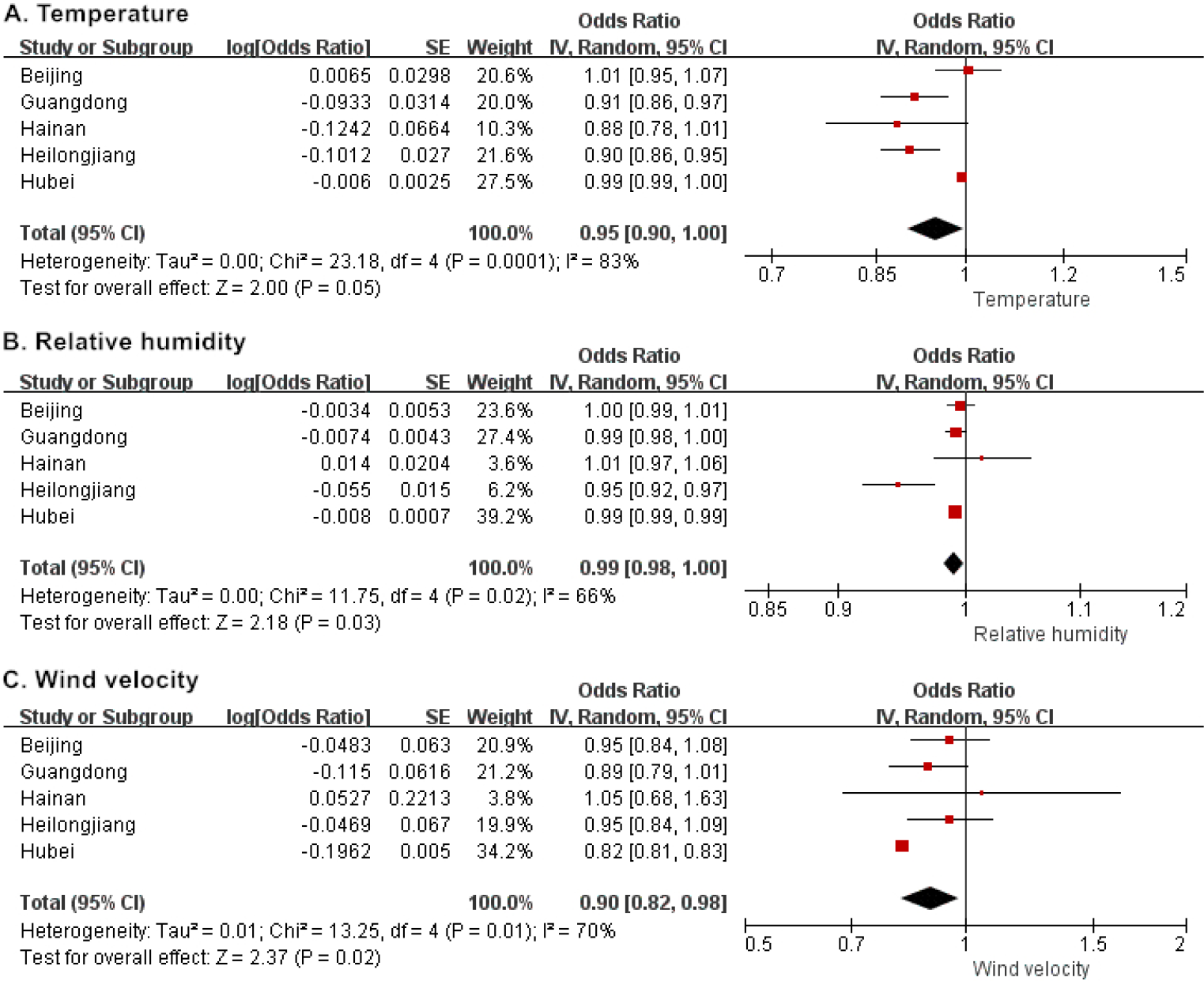
Meta-analysis of the associations between meteorological factors and COVID-19 confirmed new cases at the previous four days lag structure (lag4) in the five selected regions.

### 3.4 Exposure-response analyses

Exposure-response relationships for COVID-19 confirmed new cases associated with the continuous air pollutants (lag4) as spline functions are presented in **Figure 6**. In general, PM_2.5_ and PM_10_ were positively associated with daily confirmed new cases, and the estimates were monotonically increased with increasing concentrations. Positive associations were observed for SO_2_, NO_2_, O_3_ and CO with a certain concentration range, and reached a peak at 35 μg/m^3^, 30 μg/m^3^, 100 μg/m^3^ and 1.5 mg/m^3^, respectively. In addition, the associations became negative at lower or higher concentrations.

**Figure 6.**
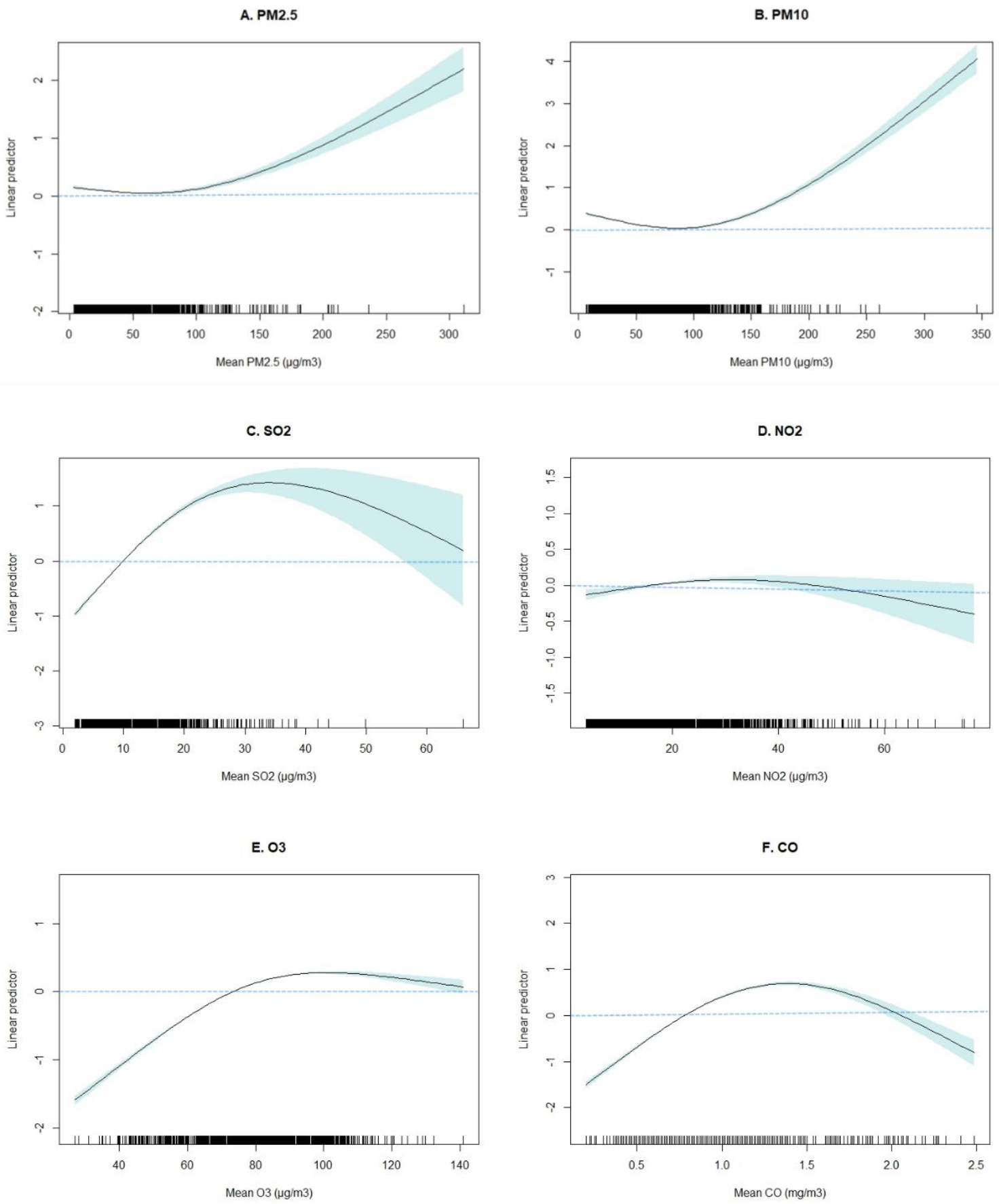
Exposure-response relationships for COVID-19 confirmed new cases with the continuous air pollutant concentrations (lag4) as spline functions.

Exposure-response relationships between COVID-19 confirmed new cases and meteorological factors (lag4) are shown in **Figure 7**. For temperature, the positive associations were observed below −10 °C and 5-20 °C, whereas the negative associations were examined ranging from −10 °C to 5 °C and above 20 °C. The positive estimates of temperature showed a peak at approximately 10 °C. The associations of relative humidity became negative and attenuated above 70%. For wind velocity, negative associations were observed ranging from 2 m/s to 5 m/s, and positive associations were observed at lower or higher speeds. For the interaction terms of meteorological factors, the number of daily COVID-19 confirmed new cases was negatively associated with the following combinations: extreme high temperature and low relative humidity; high temperature and extreme wind velocity; and extreme low relative humidity and low wind velocity.

**Figure 7.**
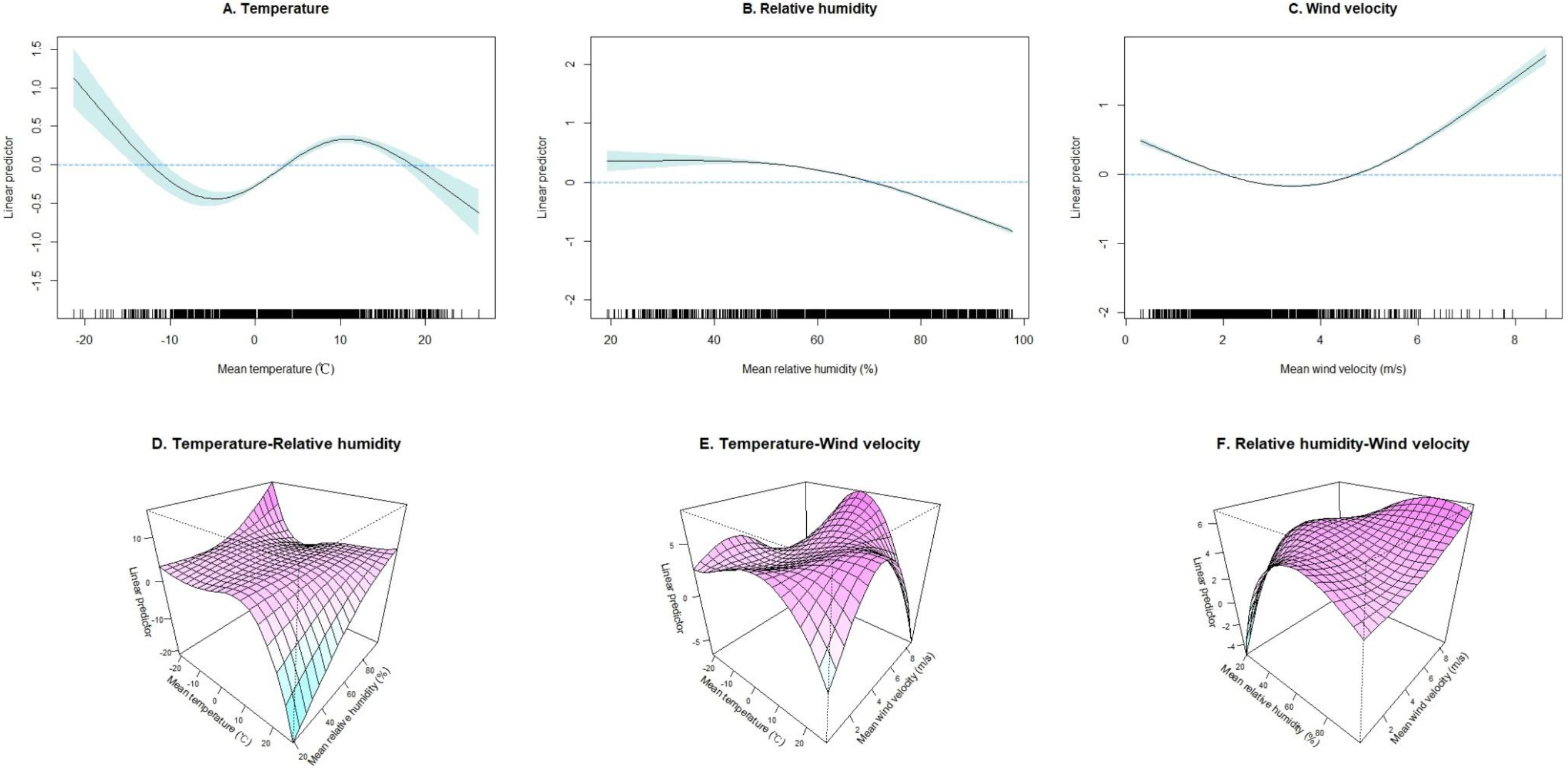
Exposure-response relationships for COVID-19 confirmed new cases with the continuous meteorological factors (lag4) as spline functions.Note: A, B and C: with the single meteorological factor. D, E and F: with interaction terms of meteorological factors.

### 3.5 Sensitivity analyses

Sensitivity analyses involved the use of four degrees of freedom in meteorological factors and variations in the level of smoothness of the time trend function (with six to eight degrees of freedom) **(Supplementary Figure A1 and Figure A2)**. All results were stable with the following exceptions: PM_2.5_ at lag05 and lag06; SO_2_ at lag06; and wind velocity at lag3.

## 4. Discussion

There were 77,578 COVID-19 confirmed cases reported in 31 Chinese provinces from January 25 to February 29, 2020. This was a time-series study to explore the associations of ambient air pollutants and meteorological factors with COVID-19 infection covering the outbreak period. Our results indicated that COVID-19 transmission may be associated with air pollutants and meteorological factors. Higher concentrations of air pollutants as well as weather with relative lower temperature, humidity and wind velocity likely favor its transmission.

Previous studies have found that respiratory infectious diseases are associated with ambient air pollutants (Gehring et al. 2020; Kan et al. 2005; Liu et al. 2019). A case-crossover study conducted in the U.S. showed that short-term exposure to elevated PM_2.5_ was associated with acute lower respiratory infections in both children and adults (Horne et al. 2018). Kan et al. also found that an increase of 10 μg/m^3^ over a 5-day moving average of PM_10_ and NO_2_ corresponded to 1.06 (1.00-1.12) and 1.22 (1.01-1.48) relative risks (RRs) of daily SARS mortality, respectively (Kan et al. 2005). However, the evidence for ambient air pollution exposure and COVID-19 infection is limited. To our knowledge, there are only two published studies that have investigated the effects of air pollutants on COVID-19 infection. Yao et al. reported significant association between NO_2_ exposure and basic reproduction number (R_0_) of COVID-19, suggesting that NO_2_ may contribute to the spread ability of COVID-19 (Yao et al. 2020a). Zhu et al. also observed positive significant associations of PM_2.5_, PM_10_, NO_2_ and O_3_ with COVID-19 confirmed cases, whereas, a negative association was observed for SO_2_ (Zhu et al. 2020). Our results indicated that higher concentrations of all air pollutants (i.e., PM_2.5_, PM_10_, SO_2_, NO_2_, O_3_ and CO) were positively associated with COVID-19 infection throughout China. Furthermore, positive significant associations of all air pollutants were also observed in Hubei, which was the province with the most serious COVID-19 infection. Regarding regional heterogeneity, the positive significant associations of PM_2.5_, PM_10_ and NO_2_ remained after the meta-analysis for the five selected regions. The pathogenic mechanisms between air pollutants and infectious diseases are still unclear. Experimental evidence has suggested that the increased transmission ability in the presence of air pollutants may not be caused by improvement of epithelial cell susceptibility to infection but may result from effects on host defenses that prevent the spread of virus (Becker and Soukup 1999). In addition, positive associations may also be affected by the relationship between population movement and virus transmission because PM_2.5_, PM_10_ and NO_2_ are traffic-related air pollutants (Yao et al. 2020a). Furthermore, we conducted exposure-response analyses between air pollutants and COVID-19 infection. The estimates of PM_2.5_ and PM_10_ monotonically increased, but, the estimates attenuated and became negative in higher concentrations of gaseous pollutants (i.e., SO_2_, NO_2_, O_3_ and CO). These results may be explained by the saturation mechanism that the underlying biochemical and cellular processes become saturated with small doses of a toxic component (Pope et al. 2009). The specific mechanisms are still unclear and need further investigation.

Meteorological factors affect both the host and virus during the outbreak of an infectious disease. On one hand, meteorological factors can influence human immunity and activity patterns (McMichael 2015). On the other hand, they can also affect the transmission and virulence of the virus (Lowen et al. 2007; Wu et al. 2016). There have been several studies investigating the effects of ambient temperature on the transmission of infectious diseases. An ecological analysis of the SARS outbreak among four Chinese cities found that the risk of increased daily SARS incidence differed greatly at high and low temperature, suggesting that temperature was a powerful indicator for the transmission of SARS-CoV (Tan et al. 2005). For COVID-19, Liu et al. found that each 1 °C increase in ambient temperature was related to 0.80 (0.75, 0.85) RR of daily confirmed case counts (Liu et al. 2020). Wang et al. and Shi et al. also examined similar associations that low temperature likely favored COVID-19 transmission (Shi et al. 2020; Wang et al. 2020). Furthermore, Shi et al. found that the RRs of the daily confirmed case counts increased to a peak at approximately 8-10 °C (Shi et al. 2020). Consistent with these results, we examined negative associations between temperature and COVID-19 confirmed new cases with a corresponding OR of 0.97 (0.97-0.98). The nonlinear exposure-response relationship indicated that negative associations were observed for temperatures ranging from −10 °C to 5 °C and above 20 °C, whereas positive associations were observed below −10 °C and 5-20 °C. The positive estimates of temperature reached a peak at approximately 10 °C, which agreed with the previous study. The overall negative estimate observed in our study may be due to the relative low temperature (5.7 ± 3.1 °C) throughout China during the study period. However, Yao et al. found that there were no significant association of COVID-19 transmission with temperature (Yao et al. 2020b). Therefore, more epidemiological and laboratory evidence for the associations of temperature are still needed.

Relative humidity is another meteorological factor that may play a part in infectious disease outbreaks. A previous study found that 12% of influenza virus transmission variability and 36% of influenza virus survival variability could be explained by relative humidity (Shaman et al. 2011). Our study found that the number of COVID-19 confirmed new cases was negatively associated with relative humidity with a corresponding OR of 0.96 (0.96-0.97). With regard to regional heterogeneity, the negative significant association remained after the meta-analysis for the five selected regions. Furthermore, the exposure-response relationship indicated that the associations became negative and attenuated above 70%, which was the mainly range of relative humidity during the study period. In line with our results, Liu et al. found that absolute humidity was significantly associated with decreased COVID-19 confirmed case counts at lag07 (RR: 0.72, 95% CI: 0.59-0.89) and lag014 (RR: 0.33, 95% CI: 0.21-0.54) (Liu et al. 2020). A previous study on SARS data also reported a significant inverse correlation between relative humidity and the number of confirmed cases in Guangzhou, Beijing and Taiyuan (Tan et al. 2005). Possible mechanism is that lower humidity in a certain range increases the stability of coronavirus and favors its transmission (Lowen et al. 2007). However, a previous study on the early COVID-19 outbreak are somewhat different from our results as it reported that the changes in temperature and humidity as spring and summer months arriving might not necessarily decline the case counts (Luo et al. 2020). Although the coming warmer season (with relative high temperature and humidity) may play an optimistic role in decreasing the transmission, the implementation of extensive public health control measures and medication mainly contribute to controlling COVID-19 epidemic (Pan et al. 2020).

Wind velocity is an important meteorological factor affecting the suspension time of pathogens in the air. High wind velocity facilitates dilution and removal of pathogens, thereby reducing the transmission potential. An ecological study found that higher daily average wind velocity corresponded to a lower secondary attack rate of SARS (Cai et al. 2007). Consistent with the previous study, we examined the negative associations between wind velocity and COVID-19 confirmed new cases in Hubei and throughout China. Regarding regional heterogeneity, the negative significant association remained after the meta-analysis for the five selected regions. The exposure-response relationship also indicated that higher wind velocity was associated with lower infection case counts when the wind velocity ranged from 2 m/s to 5 m/s. To our knowledge, we are the first to investigate the effects of wind velocity on COVID-19 transmission. Therefore, more epidemiological studies and direct laboratory evidence are needed to verify our results.

In addition, we considered the associations of meteorological factor combinations on COVID-19 infection. The three-dimensional exposure-response relationship indicated that weather with high temperature, low humidity and high wind velocity may decrease COVID-19 transmission but that weather with low temperature, low humidity and low wind velocity may favor its transmission. The survival characteristics of coronavirus support our results. An experimental study on SARS-CoV found that the virus soon lost its activity when the temperature increased to 38 °C and the relative humidity reached 95% (Chan et al. 2011). Another study on MERS-CoV, a virus also sharing genetic similarities with COVID-19, indicated that it could maintain activity for a long time both as droplets on solid surface and as aerosol in low temperature (20 °C) and low humidity (40%) conditions (van Doremalen et al. 2013). Our results were also accordance with the weather condition of COVID-19 outbreak in China and countries with similar latitude (e.g., U.S., Italy, Japan and Korea), which was in winter and early spring with relative low temperature and low humidity (Rothan and Byrareddy 2020). As summer months (hot and humid) are arriving in the Northern Hemisphere, the weather conditions may play an optimistic role in decreasing the transmission. At the same time, it should be noted that COVID-19 is pandemic throughout the world, even in several tropic countries with hot and dry weather conditions (e.g., India, Mexico and Brazil). Therefore, the implementation of extensive public health control measures (e.g., shutting down cities, extending holidays and travel ban) and medication (Pan et al. 2020) is more efficient and powerful than weather changing for controlling the COVID-19 infection.

Identifying the associations of environmental factors in different lag structures is of great importance in the estimation of health risk. Incubation period, the time of diagnosis and reporting delay should be considered in risk assessment of infectious diseases. For COVID-19, previous four days lag structure (lag4) may be the most appropriate for the OR estimations because at least five days were required for case confirmation with the shortest incubation period (three days of incubation, one day for laboratory diagnosis, and one day delay for case reporting). In addition, the associations of air pollutants and meteorological factors with COVID-19 infection at lag4 were more stable than other lag structures through sensitivity analyses. Therefore, estimates at lag4 were selected for our study.

There were several strengths in the present study. First, the study period (from January 25 to February 29, 2020) covered the COVID-19 outbreak in China, which allowed representative association estimates between environmental factors and disease infection. Second, 31 Chinese cities with diverse air pollution levels (e.g., daily mean PM_2.5_ ranging from 3.2 μg/m^3^ to 311.5 μg/m^3^) and meteorological conditions (e.g., daily mean temperature ranging from −21.3 °C to 26.3 °C) were included in our study, which provided enough span for association estimates and exposure-response analyses. Third, to our knowledge, we are the first to explore the associations of meteorological factor combinations on COVID-19 infection. The three-dimensional exposure-response relationship suggested that low temperature, low humidity and low wind velocity likely favor COVID-19 transmission.

Our study also had some limitations. First, this is a province-level ecological study without considering the implementation ability of COVID-19 control policy, urbanization rate and availability of medical resources, which may affect COVID-19 transmission and confound our results. Second, demographic variables (e.g., age, gender and history of cardiopulmonary diseases) were not considered in our study due to a deficiency of detailed information for each infectious case. Third, our study was conducted with a certain range of air pollutant concentrations and meteorological factors. The results may not be applicable to countries with different air pollution levels and weather conditions. Therefore, global studies are needed to verify our results.

## 5. Conclusions

Our study indicates that COVID-19 infection is associated with both air pollutant exposures and meteorological factors. Higher air pollutant concentrations and weather with relative lower temperature, humidity and wind velocity may favor its transmission. As summer months with increasing temperature and humidity are arriving in the Northern Hemisphere, the environmental factors and the implementation of extensive public health control measures may play an optimistic role for controlling COVID-19 epidemic.

## Data Availability

The data used in this study was public data and was derived from official websites. Therefore, all data is available in our study.

## Acknowledgements

We thank the National Health Commission for providing COVID-19 confirmed case data, the China National Environmental Monitoring Centre for providing air pollutant data, and the National Meteorological Information Center for providing meteorological data.

## Appendix A. Supplementary data

Supplementary data for this article can be found as an attached file.

## Author contributions

Han Cao: Methodology, Data curation, Formal analysis, Visualization, Writing-Original draft. Bingxiao Li, Tianlun Gu, Xiaohui Liu: Investigation, Data curation.

Kai Meng: Validation, Writing-Reviewing and Editing.

Ling Zhang: Conceptualization, Investigation, Supervision, Writing-Reviewing and Editing.

## Abbreviations

CO: carbon monoxide
IQR: interquartile range
NO_2_: nitrogen dioxide
O_3_: ozone
OR: odds ratio
PM: particulate matter
PM_10_: PM with an aerodynamic diameter ≤10 μm
PM_2.5_: PM with an aerodynamic diameter ≤2.5 μm
SO_2_: sulfur dioxide

## References

Becker S, Soukup JM. 1999. Effect of nitrogen dioxide on respiratory viral infection in airway epithelial cells. Environmental research 81:159–166, PMID: 10433848, https://doi.org/10.1006/enrs.1999.3963.

Cai QC, Lu J, Xu QF, Guo Q, Xu DZ, Sun QW, et al. 2007. Influence of meteorological factors and air pollution on the outbreak of severe acute respiratory syndrome. Public health 121:258–265, PMID: 17307207, https://doi.org/10.1016/j.puhe.2006.09.023.

Chan KH, Peiris JS, Lam SY, Poon LL, Yuen KY, Seto WH. 2011. The effects of temperature and relative humidity on the viability of the sars coronavirus. Advances in virology 2011:734690, PMID: 22312351, https://doi.org/10.1155/2011/734690.

Cui Y, Zhang ZF, Froines J, Zhao J, Wang H, Yu SZ, et al. 2003. Air pollution and case fatality of sars in the people’s republic of china: An ecologic study. Environmental health: a global access science source 2:15, PMID: 14629774, https://doi.org/10.1186/1476-069x-2-15.

Gehring U, Wijga AH, Koppelman GH, Vonk JM, Smit HA, Brunekreef B. 2020. Air pollution and the development of asthma from birth until young adulthood. The European respiratory journal, PMID: 32299858, https://doi.org/10.1183/13993003.00147-2020.

Horne BD, Joy EA, Hofmann MG, Gesteland PH, Cannon JB, Lefler JS, et al. 2018. Short-term elevation of fine particulate matter air pollution and acute lower respiratory infection. American journal of respiratory and critical care medicine 198:759–766, PMID: 29652174, https://doi.org/10.1164/rccm.201709-1883OC.

Ianevski A, Zusinaite E, Shtaida N, Kallio-Kokko H, Valkonen M, Kantele A, et al. 2019. Low temperature and low uv indexes correlated with peaks of influenza virus activity in northern europe during 2010(-)2018. Viruses 11, PMID: 30832226, https://doi.org/10.3390/v11030207.

Kan HD, Chen BH, Fu CW, Yu SZ, Mu LN. 2005. Relationship between ambient air pollution and daily mortality of sars in beijing. Biomedical and environmental sciences: BES 18:1–4, PMID: 15861770,

Li Q, Guan X, Wu P, Wang X, Zhou L, Tong Y, et al. 2020. Early transmission dynamics in wuhan, china, of novel coronavirus-infected pneumonia. The New England journal of medicine 382:1199–1207, PMID: 31995857, https://doi.org/10.1056/NEJMoa2001316.

Liu J, Zhou J, Yao J, Zhang X, Li L, Xu X, et al. 2020. Impact of meteorological factors on the covid-19 transmission: A multi-city study in china. The Science of the total environment 726:138513, PMID: 32304942, https://doi.org/10.1016/j.scitotenv.2020.138513.

Liu T, Kang M, Zhang B, Xiao J, Lin H, Zhao Y, et al. 2018. Independent and interactive effects of ambient temperature and absolute humidity on the risks of avian influenza a(h7n9) infection in china. The Science of the total environment 619-620:1358–1365, PMID: 29734613, https://doi.org/10.1016/j.scitotenv.2017.11.226.

Liu XX, Li Y, Qin G, Zhu Y, Li X, Zhang J, et al. 2019. Effects of air pollutants on occurrences of influenza-like illness and laboratory-confirmed influenza in hefei, china. International journal of biometeorology 63:51–60, PMID: 30382350, https://doi.org/10.1007/s00484-018-1633-0.

Lowen AC, Mubareka S, Steel J, Palese P. 2007. Influenza virus transmission is dependent on relative humidity and temperature. PLoS pathogens 3:1470–1476, PMID: 17953482, https://doi.org/10.1371/journal.ppat.0030151.

Lu R, Zhao X, Li J, Niu P, Yang B, Wu H, et al. 2020. Genomic characterisation and epidemiology of 2019 novel coronavirus: Implications for virus origins and receptor binding. Lancet 395:565–574, PMID: 32007145, https://doi.org/10.1016/s0140-6736(20)30251-8.

Luo W, Majumder MS, Liu D, Poirier C, Mandl KD, Lipsitch M, et al. 2020. The role of absolute humidity on transmission rates of the covid-19 outbreak. medRxiv:2020.2002.2012.20022467, PMID https://doi.org/10.1101/2020.02.12.20022467.

McMichael C. 2015. Climate change-related migration and infectious disease. Virulence 6:548–553, PMID: 26151221, https://doi.org/10.1080/21505594.2015.1021539.

Nhung NTT, Schindler C, Dien TM, Probst-Hensch N, Perez L, Kunzli N. 2018. Acute effects of ambient air pollution on lower respiratory infections in hanoi children: An eight-year time series study. Environment international 110:139–148, PMID: 29128032, https://doi.org/10.1016/j.envint.2017.10.024.

Pan A, Liu L, Wang C, Guo H, Hao X, Wang Q, et al. 2020. Association of public health interventions with the epidemiology of the covid-19 outbreak in wuhan, china. Jama, PMID: 32275295, https://doi.org/10.1001/jama.2020.6130.

Peci A, Winter AL, Li Y, Gnaneshan S, Liu J, Mubareka S, et al. 2019. Effects of absolute humidity, relative humidity, temperature, and wind speed on influenza activity in toronto, ontario, canada. Applied and environmental microbiology 85, PMID: 30610079, https://doi.org/10.1128/aem.02426-18.

Pope CA, 3rd, Burnett RT, Krewski D, Jerrett M, Shi Y, Calle EE, et al. 2009. Cardiovascular mortality and exposure to airborne fine particulate matter and cigarette smoke: Shape of the exposure-response relationship. Circulation 120:941–948, PMID: 19720932, https://doi.org/10.1161/circulationaha.109.857888.

Rothan HA, Byrareddy SN. 2020. The epidemiology and pathogenesis of coronavirus disease (covid-19) outbreak. Journal of autoimmunity 109:102433, PMID: 32113704, https://doi.org/10.1016/j.jaut.2020.102433.

Samet JM, Dominici F, Zeger SL, Schwartz J, Dockery DW. 2000. The national morbidity, mortality, and air pollution study. Part i: Methods and methodologic issues. Research report (Health Effects Institute):5-14; discussion 75–84, PMID: 11098531,

Shaman J, Goldstein E, Lipsitch M. 2011. Absolute humidity and pandemic versus epidemic influenza. Am J Epidemiol 173:127–135, PMID: 21081646, https://doi.org/10.1093/aje/kwq347.

Shi P, Dong Y, Yan H, Zhao C, Li X, Liu W, et al. 2020. Impact of temperature on the dynamics of the covid-19 outbreak in china. The Science of the total environment 728:138890, PMID: 32339844, https://doi.org/10.1016/j.scitotenv.2020.138890.

Tan J, Mu L, Huang J, Yu S, Chen B, Yin J. 2005. An initial investigation of the association between the sars outbreak and weather: With the view of the environmental temperature and its variation. Journal of epidemiology and community health 59:186–192, PMID: 15709076, https://doi.org/10.1136/jech.2004.020180.

van Doremalen N, Bushmaker T, Munster VJ. 2013. Stability of middle east respiratory syndrome coronavirus (mers-cov) under different environmental conditions. Euro surveillance: bulletin Europeen sur les maladies transmissibles = European communicable disease bulletin 18, PMID: 24084338, https://doi.org/10.2807/1560-7917.es2013.18.38.20590.

Wang M, Jiang A, Gong L, Luo L, Guo W, Li C, et al. 2020. Temperature significant change covid-19 transmission in 429 cities. medRxiv:2020.2002.2022.20025791, PMID: https://doi.org/10.1101/2020.02.22.20025791.

Wu X, Lu Y, Zhou S, Chen L, Xu B. 2016. Impact of climate change on human infectious diseases: Empirical evidence and human adaptation. Environment international 86:14–23, PMID: 26479830, https://doi.org/10.1016/j.envint.2015.09.007.

Yao Y, Pan J, Liu Z, Meng X, Wang W, Kan H, et al. 2020a. Ambient nitrogen dioxide pollution and spread ability of covid-19 in chinese cities. medRxiv:2020.2003.2031.20048595, PMID: https://doi.org/10.1101/2020.03.31.20048595.

Yao Y, Pan J, Liu Z, Meng X, Wang W, Kan H, et al. 2020b. No association of covid-19 transmission with temperature or uv radiation in chinese cities. European Respiratory Journal:2000517, PMID: https://doi.org/10.1183/13993003.00517-2020.

Zhou F, Yu T, Du R, Fan G, Liu Y, Liu Z, et al. 2020. Clinical course and risk factors for mortality of adult inpatients with covid-19 in wuhan, china: A retrospective cohort study. Lancet 395:1054–1062, PMID: 32171076, https://doi.org/10.1016/s0140-6736(20)30566-3.

Zhu Y, Xie J, Huang F, Cao L. 2020. Association between short-term exposure to air pollution and covid-19 infection: Evidence from china. The Science of the total environment 727:138704, PMID: 32315904, https://doi.org/10.1016/j.scitotenv.2020.138704.

